# Mapping the last mile: micro-stratification for sustained visceral leishmaniasis elimination in Bangladesh

**DOI:** 10.1101/2025.08.26.25334417

**Authors:** Shomik Maruf, Md Rasel Uddin, Farhana Rahman Luba, Soumik Kha Sagar, Debashis Ghosh, Sakhawat Hossain, Megha Raj Banjara, Axel Kroeger, Christine Halleux, Abraham Aseffa, Dinesh Mondal

## Abstract

**Background:** Bangladesh became the first country to achieve World Health Organization (WHO) validation for eliminating visceral leishmaniasis (VL, Kala-azar) as a public-health problem in 2023. Sustaining this milestone demands a post-validation surveillance strategy that concentrates its efforts on residual transmission foci and deploys resources efficiently. We therefore conducted the country’s first Mouza-level micro-stratification to refine risk maps and guide targeted interventions.

**Methods:** The study used routinely reported VL line-list data (2017-2022) from the national DHIS2 platform for every Upazila that recorded ≥1 VL case. Each Mouza—the smallest administrative unit—was categorised as high (≥3 new VL cases), moderate (2 cases), low (1 case) or non-endemic (0 cases) over the six-year period. Hot-spot maps were created in Python. Associations between endemicity (endemic vs non-endemic) and mean annual climate variables were tested with chi-squared statistics, yielding odds ratios (OR) with 95% confidence intervals (CI). Historical sand-fly density data from 51 purposively sampled Mouzas were compared across endemicity strata using one-way ANOVA. Among 15,982 Mouzas in 119 case-reporting Upazilas, only 428 (2.7%) reported ≥1 new VL case between 2017 and 2022. High-endemic Mouzas (n = 29; 0.18 %) accounted for 36% of total incident cases and clustered primarily in Mymensingh, Dhaka and Rajshahi divisions. However, year-on-year mapping showed contraction of endemic Mouzas with no sustained new foci.

**Conclusions:** VL transmission in Bangladesh is now intensely focal, confined to <3% of Mouzas within historically endemic Upazilas. Restricting surveillance, IRS and active case detection through micro-stratification of endemic areas could largely cut vector-control costs while safeguarding elimination. Periodic updating Mouza-level risk maps and integrating high-resolution climate and entomological data will be essential to prevent resurgence and can serve as a model for other countries nearing VL elimination.

**Author Summary:** We studied visceral leishmaniasis (also called kala-azar), a parasitic disease spread by sand flies that can be deadly if not treated. Since Bangladesh met global targets for public health elimination of this disease in 2023, the key challenge now is to keep it from coming back. We asked a simple question: where, exactly, do cases still occur? Using routine records from the national health information system (2017–2022), we created detailed maps at the level of mouza (small villages or village clusters). We found that recent illness was highly concentrated. Only about 3% of villages reported any new case over six years, and fewer than thirty villages held more than one-third of all cases. This pattern tells us the disease now survives in small pockets rather than everywhere. By pointing health teams to these pockets, our maps can help focus actions such as going door-to-door to find patients early, supporting their care, and controlling sand flies in the right places. We share a practical approach that can guide local planning in Bangladesh and can be adapted in other countries working to sustain control.

## Introduction

Visceral Leishmaniasis (VL), often known as Kala-azar (KA), is a potentially life-threatening disease caused by Leishmania parasites and transmitted by the bites of female phlebotomine sandflies [1]. An approximate number of 50,000 to 90,000 new cases of VL occur globally every year [2]. Historically, densely populated and impoverished countries like India, Bangladesh, and Nepal in the Indian Subcontinent (ISC) had contributed substantially to VL burden [3–4]. These three countries committed to eliminating VL by 2015 through a memorandum of understanding (MoU) signed in 2005 [1]. While Nepal successfully reached the elimination goals in 2013, it unfortunately lost this status by 2017, highlighting the challenges of sustaining such achievements [5]. On a similar note, Bangladesh became the first country to receive official validation from the World Health Organization (WHO) in 2023 for eliminating visceral leishmaniasis as a public health problem. However, sustainment of this elimination status is crucial to ensure the success of the Regional Kala-azar Elimination Initiative. Recognizing the importance of sustained elimination, the National Kala-azar Elimination Programme (NKEP) in Bangladesh is currently prioritizing effective surveillance as part of its post-validation efforts [6].

The efforts encompass investigations into strategies for optimizing targeted risk surveillance, assessing disease burden, and implementing integrated intervention programs [7]. The sub-district (Upazila), also known as Upazila, serves as the primary reporting unit for this surveillance system under the NKEP [8]. Upazila is considered a focal point of the government administrative unit of Bangladesh, consisting of several Mouzas with an average population of around 333,653 and a mean area of 298 square kilometres [9–11]. According to DHIS2 data of 2019, 100 Upazilas among 495 were considered endemic for VL, further categorized as low, moderate, and high endemic regions [8]. However, the epidemiology of visceral leishmaniasis (VL) has undergone substantial changes in recent years, particularly during the consolidation phase. In addition to the emergence of VL in new areas, a concerning resurgence of cases was observed in areas where the disease was once prevalent [12]. Moreover, a continuous allocation of resources to cover a larger sub-district (Upazila) area for disease surveillance and vector control has become a major concern for the NKEP. Given these factors, directing concentrated attention towards specific Mouzas, the base-level administrative unit consisting of subplots of Upazilas, has become an essential strategy [13,14]. They have an average area of 2.5 square kilometres and a population of 2,806 [15].

An analysis of historical VL data at the Upazila level reveals that the entire population is not equally at risk, as the disease disproportionately affects the poorest and most marginalized communities. While the Upazila-wide approach was practical during the attack phase, it clearly requires revision in the post-elimination phase. The limited resources available in Bangladesh necessitate a more targeted approach to VL control. Implementing interventions across large populations covering vast areas of the endemic Upazilas can be inefficient, time-consuming, and may not yield optimal results. To maximize the resource efficiency in the post-validation phase, a shift towards focal surveillance within smaller geographical units than the Upazila is essential. Focusing on smaller areas or micro-stratification will lessen the burden of the NKEP in this regard. Therefore, in this study, we have analysed spatial data to investigate VL risk at the Mouza level by assessing the incidence of New VL cases in Bangladesh between 2017 and 2022. We have also assessed the disease shifting patterns during this period and ascertained the probable causes. This strategic information will be helpful to the NKEP in identifying VL hotspots and new foci in Bangladesh and plan targeted interventions in high-risk active foci areas, ensuring the optimum use of the available resources to sustain the elimination of VL in Bangladesh.

## Methods

### Study design

This study was the first-ever micro-stratified study of visceral leishmaniasis (VL), aiming to refine our understanding of VL endemicity and its evolving disease patterns within Bangladesh. We conducted the micro-stratification using existing secondary data available at the Directorate General of Health Services (DGHS), Bangladesh. The stratification was done at the Mouza level within the Upazila utilizing the data available from the line listing of VL cases, categorized as high, moderate, low, and non-endemic Mouzas.

### Questionnaire/checklist design

We developed specific forms to collect secondary data at the Mouza level. VL burden data was collected from the DGHS Bangladesh, which has a line listing of VL. The Mouza-level information was collected using the questionnaire template, which had two parts; the first part contains demographic, climate, and vector density data, whereas the second part contains VL disease, diagnosis and treatment data. These tools were finalized following the technical consultation meetings with NKEP and other stakeholders.

### Study/mapping unit

This study selected Mouzas as the unit for micro-stratification as they are the smallest administrative units of communities living in Bangladesh. Each Mouza in the 495 Upazila was stratified into high, moderate, low, and no VL risk Mouzas. The endemicity was determined after reviewing the previous six-year VL data (2017-2022) available at the DGHS Bangladesh.

### Data Collection

Mouza-wise VL burden data from all the Upazilas that reported any new VL cases during the period of 2017-2022 was obtained from the DGHS Bangladesh. New VL cases were defined as the National Kala-azar Management Guideline [1]. The lists of Mouzas under the Upazilas were finalized and all the Mouzas were categorized as per their endemicity, based on the review of the VL burden data obtained from DGHS. The Mouzas that reported at least one new VL case during this period were considered as VL endemic Mouzas, and rest were considered as VL non-endemic Mouzas (Fig 1). At first, we plotted year-wise VL non-endemic and endemic Mouzas into maps and observed shifting pattern for VL endemicity. Similarly, an endemicity grading was done and a hotspot Mouza-wise map was developed based on a six-year cumulative VL case burden. The VL endemic Mouzas were sub-categorized into low, moderate and high VL endemic Mouzas based on the number of cases reported (Table 1).

**Fig 1:**
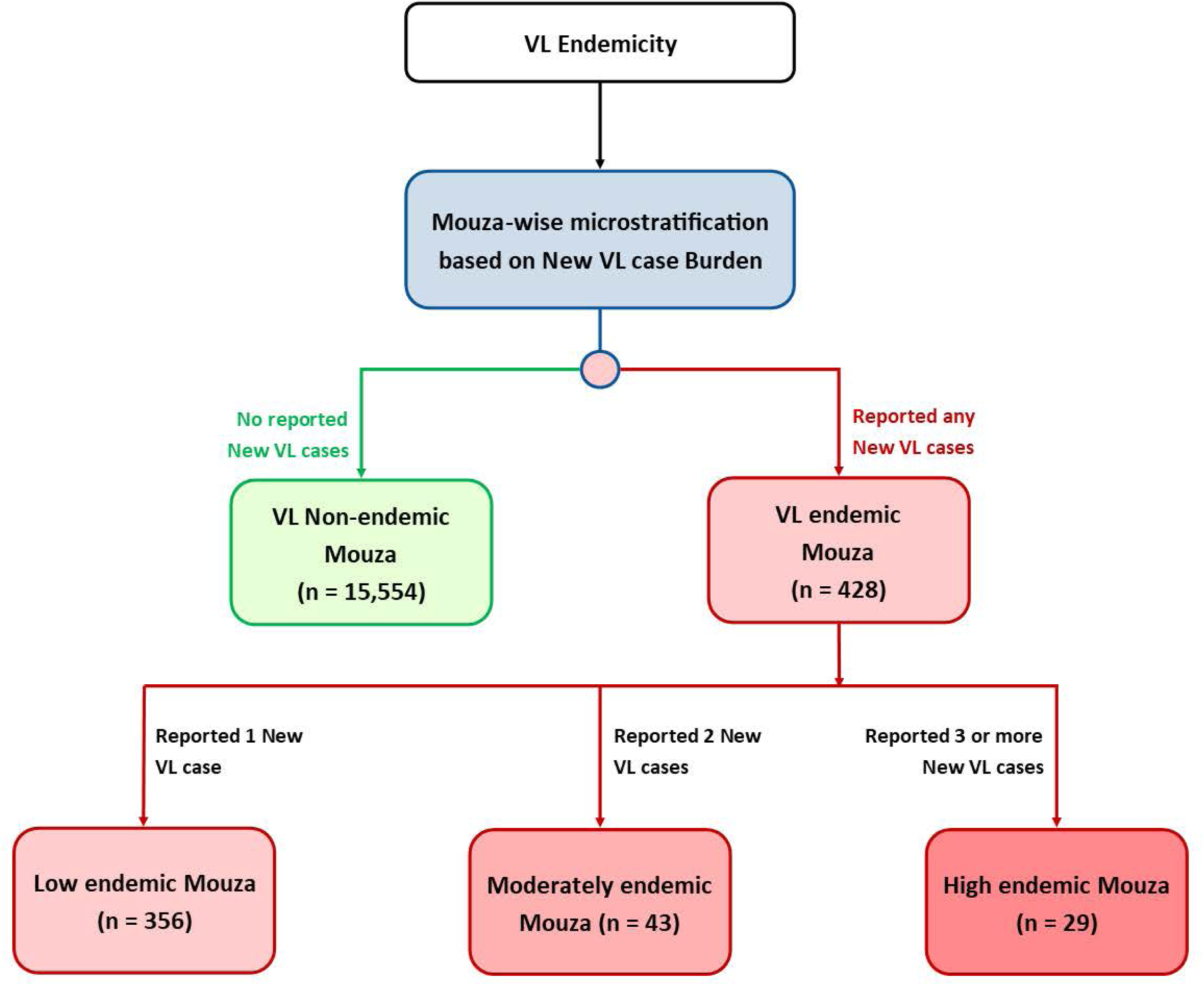
Algorithm for micro-stratification of VL endemicity.

**Table 1.**
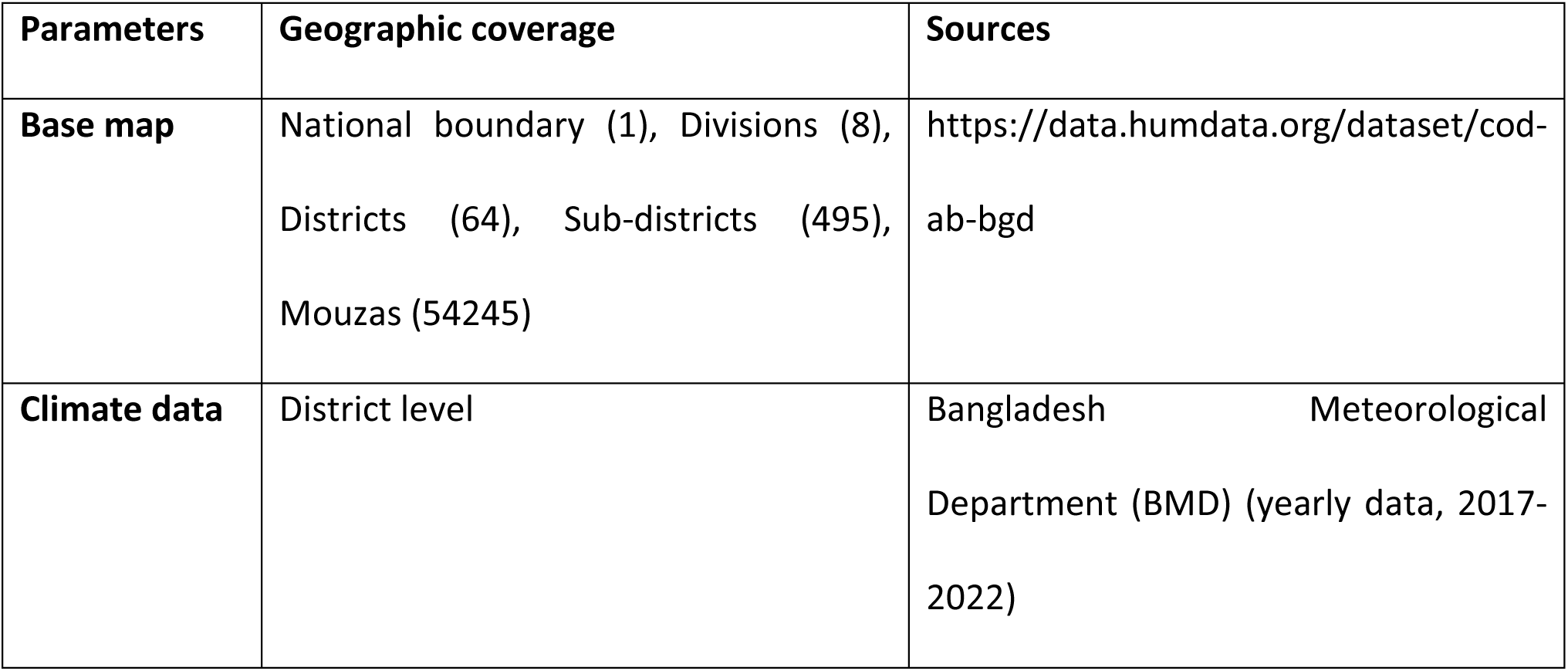
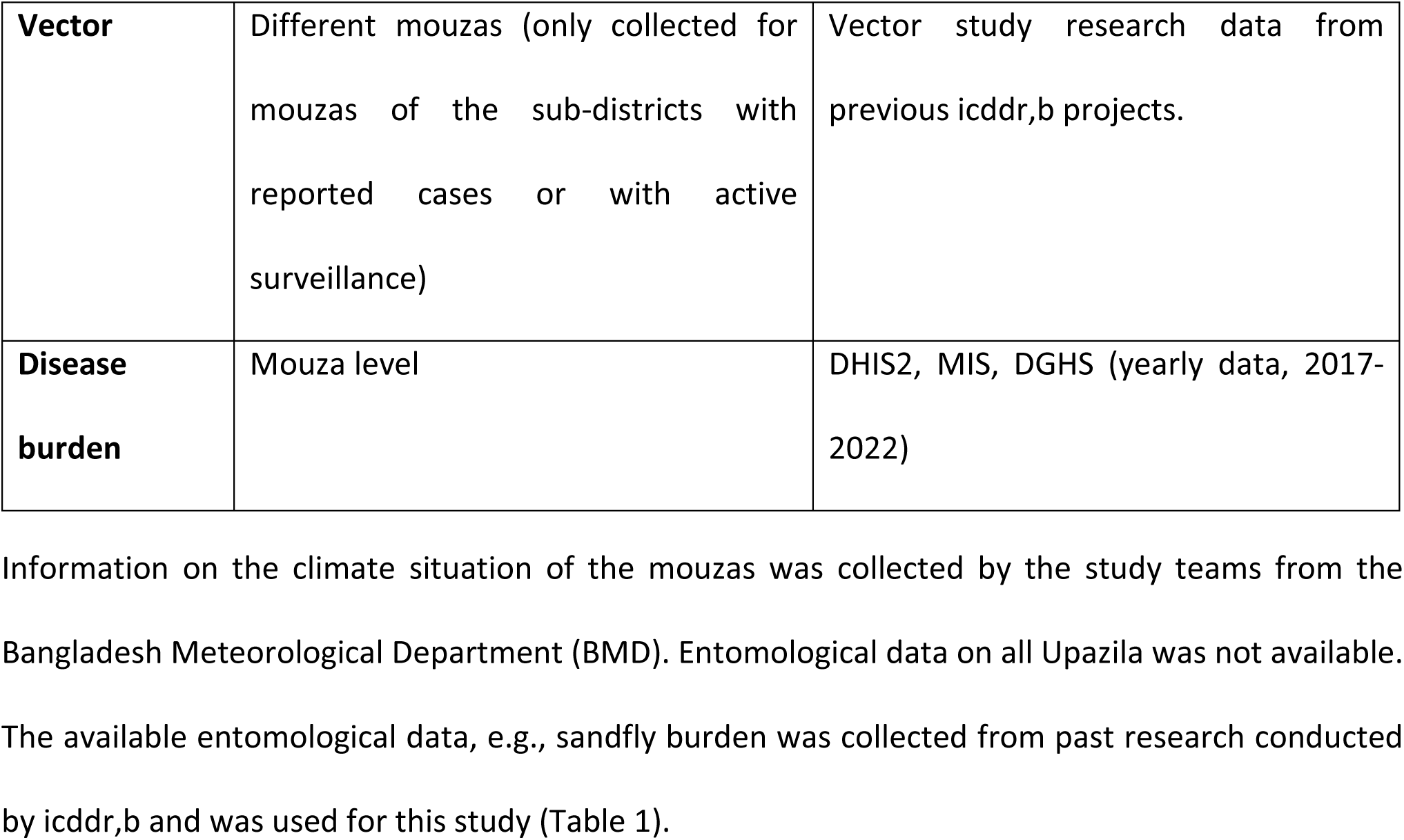
Different parameters and their sources of information.

| Parameters | Geographic coverage | Sources |
| --- | --- | --- |
| Base map | National boundary (1), Divisions (8),<br>Districts (64), Sub-districts (495),<br>Mouzas (54245) | https://data.humdata.org/dataset/cod-ab-bgd |
| Climate data | District level | Bangladesh Meteorological<br>Department (BMD) (yearly data, 2017-2022) |
| **Vector** | Different mouzas (only collected for mouzas of the sub-districts with reported cases or with active surveillance) | Vector study research data from previous icddr,b projects. |
| **Disease burden** | Mouza level | DHIS2, MIS, DGHS (yearly data, 2017-2022) |

### Data management and analysis

Data were extracted from DHIS2, MIS, DGHS in electronic format, which were cross-checked and verified for any discrepancies. A meticulously crafted data entry program was established using Microsoft Access 2019, ensuring a robust foundation for data integrity. Before input into this system, we implemented a rigorous double-checking process, strictly adhering to data management guidelines. The subsequent dataset analysis was conducted utilizing STATA (Stata Statistical Software: Release 16, College Station, Texas, USA: StataCorp LLC). Descriptive statistics were generated to unveil the inherent characteristics of the data. Parametric methods were selectively employed based on the distributional attributes of variables for comparing mean differences. ANOVA was employed to compare mean differences between groups, providing insight into the dataset. This meticulous approach ensured the fidelity of the data analysis process and fortified the reliability of the study’s findings. Chi-squared tests were performed to compare between groups and odds ratio was calculated to determine association of endemicity with different parameters. All the tests were two-tailed, and a p-value of <0.05 was considered statistically significant. Python (version 3.12.0) was used to generate hotspot maps.

## Results

We determined the endemicity of the Mouzas for each year from 2017 to 2022 and plotted them in the map. The map over the duration showed decline in the number of endemic Mouzas (Table 1), but no shifting pattern of the endemicity or no new foci for VL cases (Fig 2).

**Fig 2:**
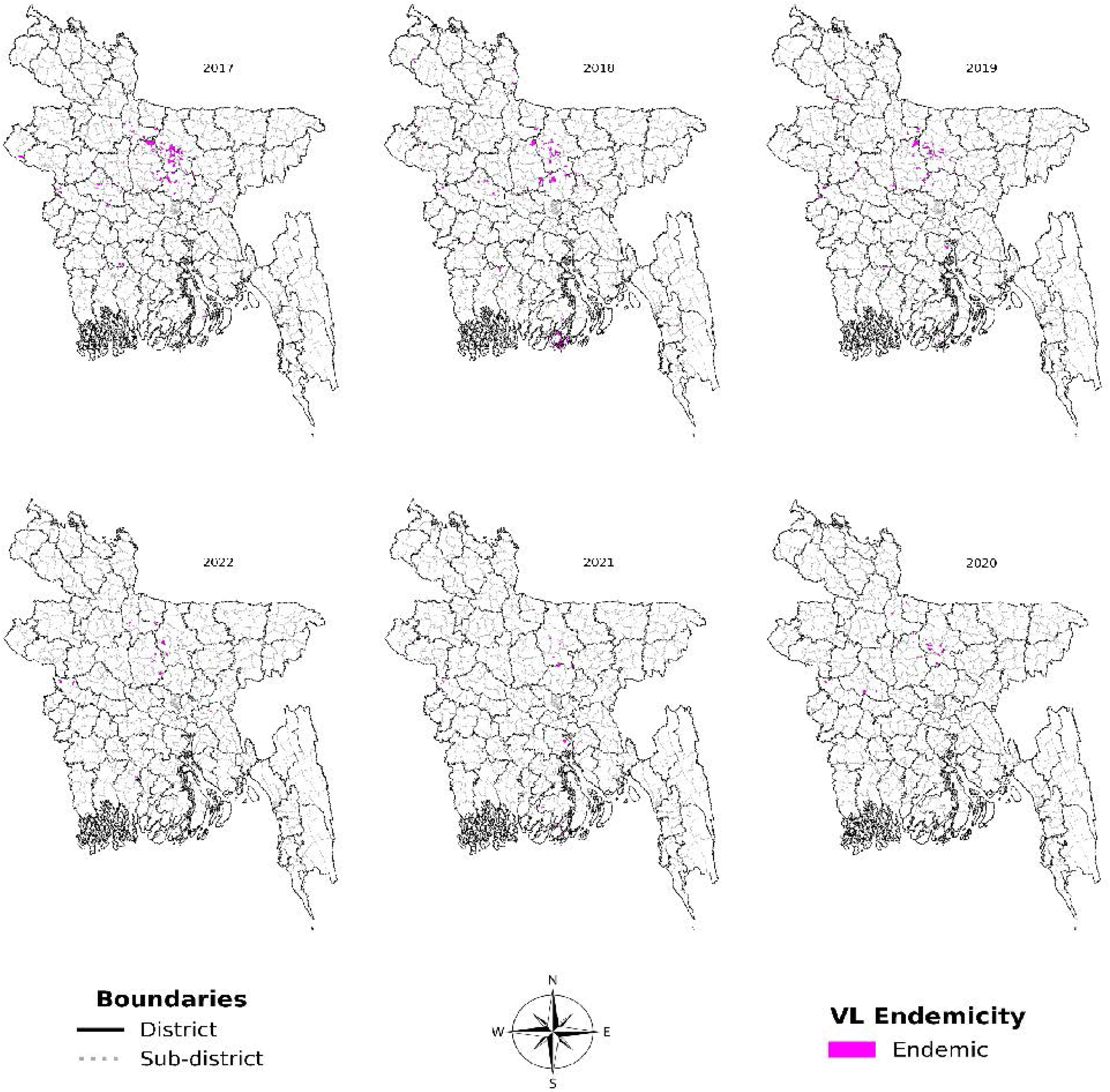
Geographical Distribution of Mouza-wise VL endemicity from 2017-2022 (clockwise).

**Fig 3:**
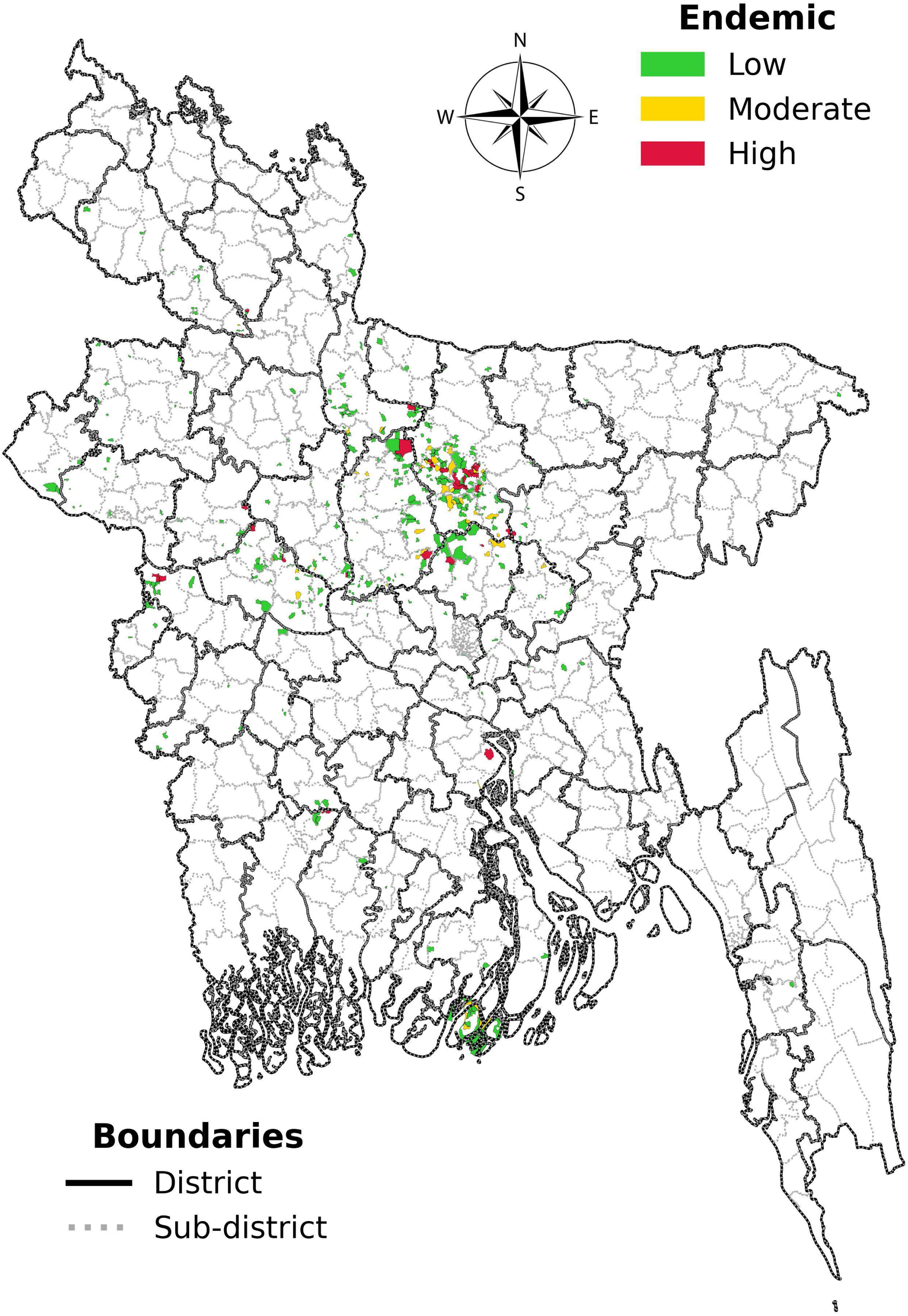
Mouza-wise VL hotspot mapping based on number of reported new cases between 2017-2022.

We further performed the same exercise based on the cumulative burden of New VL cases. We have found that 428 Mouzas reported at least one new VL case – which we defined as VL endemic Mouzas. These endemic mouzas belong to 119 endemic sub-districts (Upazila) from 39 districts from all eight divisions of the country. The endemic Mouzas were further categorized based on the incidence of New VL cases in last six years into low (n = 356), moderate (n = 243) and high endemic (n = 29) Mouzas with 1, 2 and ≥3 New VL cases reported, respectively (Table 2).

**Table 1:** Yearly distribution of endemic Mouzas for VL.

| Year | VL Endemic Mouza |
| --- | --- |
| 2017 | 176 |
| 2018 | 117 |
| 2019 | 92 |
| 2020 | 45 |
| 2021 | 33 |
| 2022 | 45 |

**Table 2:** Category of Mouzas with reported leishmanial cases between 2017-2022. The total number of Mouzas in these 119 Upazilas is 15982, whereas only 428 Mouzas reported new VL cases, which is about 2.7% of the total Mouza count in those Upazila (Table 2). We plotted all the 428 endemic Mouzas in the Mouza-wise map. We observed large clustering of endemic Mouzas in the Mymensingh (35.28%), Dhaka (28.74%), and Rajshahi division (21.50%). We found that the highest number of endemic Mouzas for VL was in Mymensingh district, followed by Tangail and Pabna districts encompassing 115 (26.9%), 66 (15.4%) and 40 (9.4%) endemic mouzas, respectively (S1 Table). Rest of the endemic Mouzas has been distributed sporadically in different parts of the country, especially along the western borders with India and a small cluster in the southern part of Barishal (Fig 3).

| Mouza Type | No. of New VL case | No. of Mouzas (N = 15982), n (%) |
| --- | --- | --- |
| Non-endemic | 0 | 15554 (97.3) |
| Endemic | $\geq 1$ | 428 (2.7) |
| · Low endemic | 1 | 356 (2.2) |
| · Moderate endemic | 2 | 43 (0.3) |
| · High endemic | $\geq 3$ | 29 (0.2) |

Endemic Mouzas were significantly more likely to lie in less warm (< 31°C) and less sun-exposed (<6 hour sunlight day) micro-climates (OR = 2 and 1.6, respectively). Paradoxically, household sand-fly counts were higher in non-endemic Mouzas (54 ± 58 flies night) than in endemic ones (9 ± 24; p < 0.001), a pattern that may reflect intensive post-2018 IRS concentrated in the few remaining transmission hotspots (see supporting information).

## Discussion

Bangladesh has achieved public health elimination for VL in 2023 as the first country in the world. This remarkable achievement has propelled the National Kala-azar Elimination Programme (NKEP) to set two more targets – firstly, zero transmission of infection by 2025 and secondly, zero case reporting by 2030. However, to sustain the current success and march towards achieving the other two, the programme needs to devise a strategy through optimization of its available resources.

Periodic surveillance for new cases along with targeted vector control measures is imperative in the VL endemic areas to sustain VL elimination. To address this issue, we have tried to micro-stratify the VL endemic Upazilas into endemic Mouzas. Here in this study, we have tried to re-adjust our focus taking Mouza – the smallest administrative unit in Bangladesh as an implementation unit for VL control rather than Upazilas in the post-validation phase. The study revealed that a total of 119 Upazilas reported at least one new VL case between the period of 2017 – 2022. The total number of Mouzas in these 119 Upazilas is 15,982 – out of which only 428 (2.7%) Mouzas reported new VL cases. This nationwide Mouza-level analysis confirms that VL transmission has contracted into a handful of residual micro-foci since Bangladesh achieved the WHO elimination target in 2017. The pattern emulates the “last-mile” heterogeneity seen for malaria in Sri Lanka, lymphatic filariasis in Nigeria and VL in India, underscoring a well-established epidemiological principle that, transmission becomes increasingly focal with the reduction of incidence to zero [18–20].

With <3 % of Mouzas still reporting new VL, blanket Upazila-wide interventions will no longer be cost-effective. Redirecting IRS and active case detection to these endemic Mouzas alone could cut annual vector-control expenditure by 80–90 % (based on a current cost of US$ 34-38 per protected household) per year without jeopardising impact, ensuring maximum utilization of available resources [21–22]. Additionally, DHIS2’s configurable dashboards are widely used for national disease surveillance and already powering DGHS hotspot dashboards. This can likewise be set to refresh at regular intervals (quarterly/yearly) at Mouza scale, instantly flagging areas that require a rapid test– treat–track response.

The distribution of cases on the hotspot mapping shows clustering of cases in the northern part of the country mostly comprising of Mymensingh and Jamalpur districts of Mymensingh division and Tangail and Gazipur districts of Dhaka division. We have also noticed a small cluster in the western part of the Padma River (Pabna, Sirajganj and Naogaon districts of Rajshahi division) and another in the southern part of the country mainly in the Patuakhali district of Barishal division. Rest of the cases are scattered sporadically in different parts of the country.

Various studies previously showed roles of vector density with the endemicity of an area for VL [23]. However, in our study we observed a contrasting situation with significantly higher sandflies collected from the households in VL non-endemic areas compared to VL endemic areas (see supporting information). The sharp decline in sandfly density from 2018 and onwards can be attributed to nationwide insecticide residual spraying (IRS) in all the VL endemic Upazila between 2015-2017. However, since 2018 more focussed IRS activity was performed targeting areas from where new VL cases were coming. Moreover, COVID-19 played a vital role in the disruption of regular IRS activities, especially in areas with no new VL cases in the previous years. Both these factors may have contributed to the sharp rise in of sandfly density in the VL non-endemic areas in 2020. Furthermore, we could only obtain vector density data for only 51 (nine VL non-endemic and forty-two VL endemic) of 15,982 Mouzas (<0.4 %), collected from various research projects with heterogeneous trapping methods and sampling seasons which was inadequate to draw any inference on the impact of vector density on endemicity.

Climate change has been mentioned to have pivotal impacts on the transmission and spread of infectious diseases, particularly vector-borne diseases (VBDs) such as visceral leishmaniasis (VL) [16]. Multiple studies in the past have found strong correlation between endemicity of an area with its climate factors, such as, average temperature, humidity, rainfall [17]. Unfortunately, we could only find climate variables (temperature, rainfall, sunshine duration, humidity) at district or, at best, Upazila scale. Therefore, differences between Mouzas within same districts could not be recorded. Moreover, we could not explore short-term weather anomalies (e.g., post-monsoon humidity spikes) that are known to influence Phlebotomus dynamics and VL transmission risk.

Despite the lack of evidence on the probable associating factors for VL endemicity, it is clear that at the post elimination stage the cases are much sparser with only a handful of Mouzas from the endemic Upazilas are still reporting new cases. Moreover, reporting of new VL cases outside the listed hundred endemic Upazilas warrants attention as well. Following the current strategy, addressing the need for a periodic surveillance and vector control measures in all these Upazilas will be a gigantic task requiring substantial amount of resources. However, concentrating the similar efforts on a micro level can ensure maximum utilization of available resources without the need for increasing the cost for additional resources. Studies on various vector borne diseases in Asia and Africa on micro-stratification have proven to devise a more targeted and effective allocation of public health resources [24–27]. Bangladesh, being the pioneer in eliminating visceral leishmaniasis as a public health problem thus must strive to create an example for other countries closing in on VL elimination on how to sustain the success in the post elimination phase. A periodic update of the micro-stratification of VL endemic Mouzas aligning with the five-yearly operational plan of NKEP can thus be an excellent strategy for Bangladesh and other countries nearing to their public health elimination of VL.

### Limitation

The absence of comprehensive entomological data informing routine, longitudinal sand-fly monitoring limited the ability to correlate year-on-year changes in vector abundance with shifting VL incidence. Mouza-wise climate variables were not available which was insufficient to accurately capture localized environmental influences on VL endemicity. Key demographic and geographical modifiers of VL risk, e.g., housing material, livestock corrals, sleeping habits, income level, etc. were unavailable at Mouza level, potentially excluding important explanatory variables. Therefore, the microstratification analysis ultimately only used the data on new cases and vector presence. The model would have been more refined had we been able to identify additional potential contributing factor for which data was available at Mouza level. Finally, Cost-savings projections relied on national unit-cost standards assuming perfect targeting and operational efficiency. A real-world savings may be lower once mobilization, logistics, and community-engagement costs for micro-foci are included.

## Conclusion

This study highlights the potential to consider a strategic pivot from blanket Upazila coverage to micro-stratification at the Mouza level during the post-validation period of VL elimination for the National for Kala-azar Elimination Programme in Bangladesh. Despite the lack of various key denominators, the findings suggest that focusing on smaller administrative units rather than entire Upazilas will enhance surveillance efficiency, optimizes resource use, and sustains VL elimination efforts. Periodic updates to the micro-stratification framework with inclusion of high-resolution climate and entomological data will be essential to ensure continued success in controlling VL in Bangladesh.

## Ethical Consideration

This research is based on secondary data obtained from DHIS2, Management Information System (MIS), Directorate General of Health Servies (DGHS). The study was approved by the icddr,b Ethical Review Committee (PR-23016).

## Data availability

The data underlying the results presented in this study will be provided by the corresponding author upon request.

## Acknowledgement

We thank National Kala-azar Elimination Programme (NKEP) for sharing relevant data on VL burden in Bangladesh.

## Author contributions

All authors contributed to the work in accordance with ICMJE criteria, including major contributions to the study’s conception, data acquisition, analysis, interpretation, drafting, or critical revision of the manuscript. Each author has given final approval for the version to be published and is responsible for the accuracy and integrity of their respective contributions.

## Supporting information

**S1 Table:**
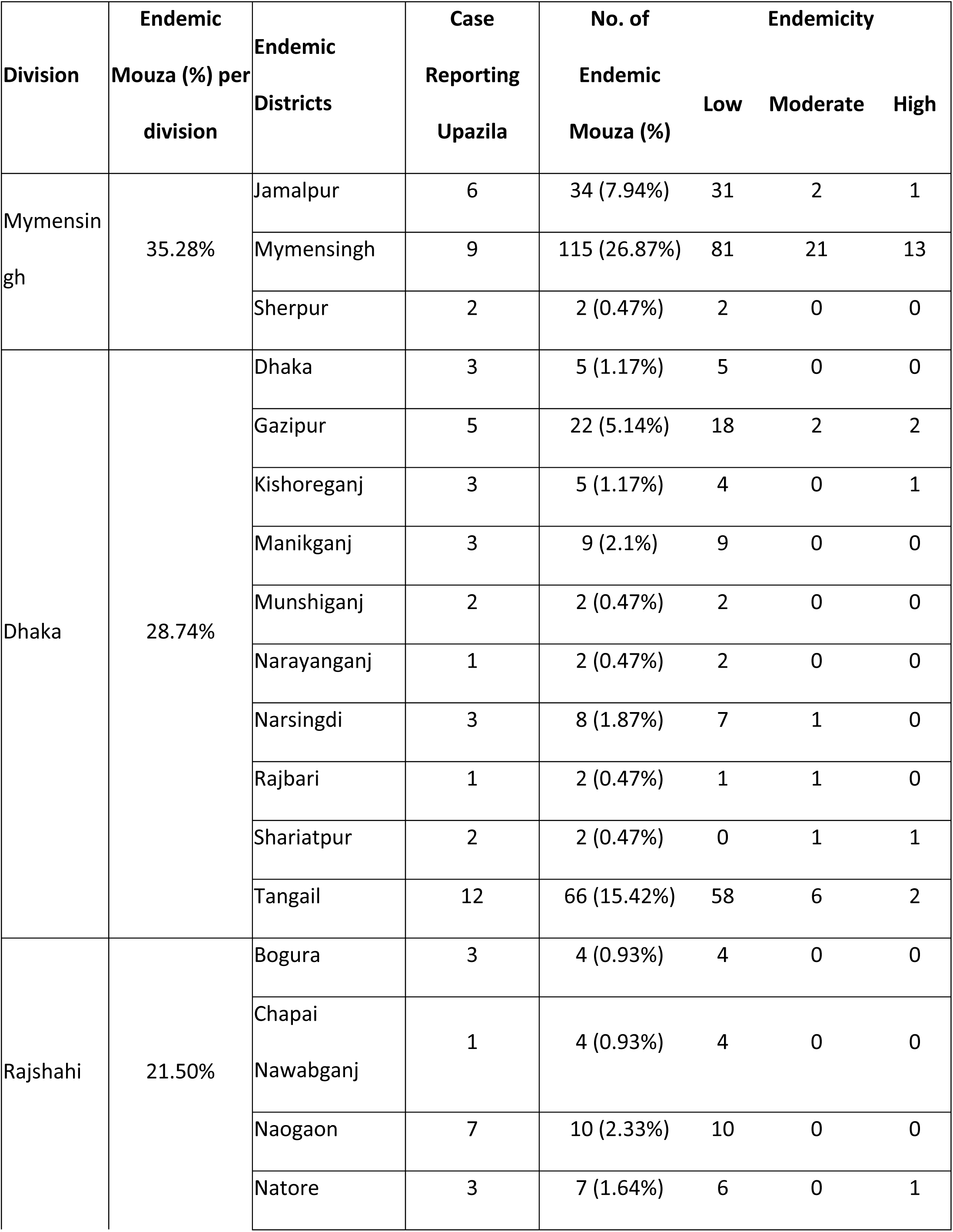
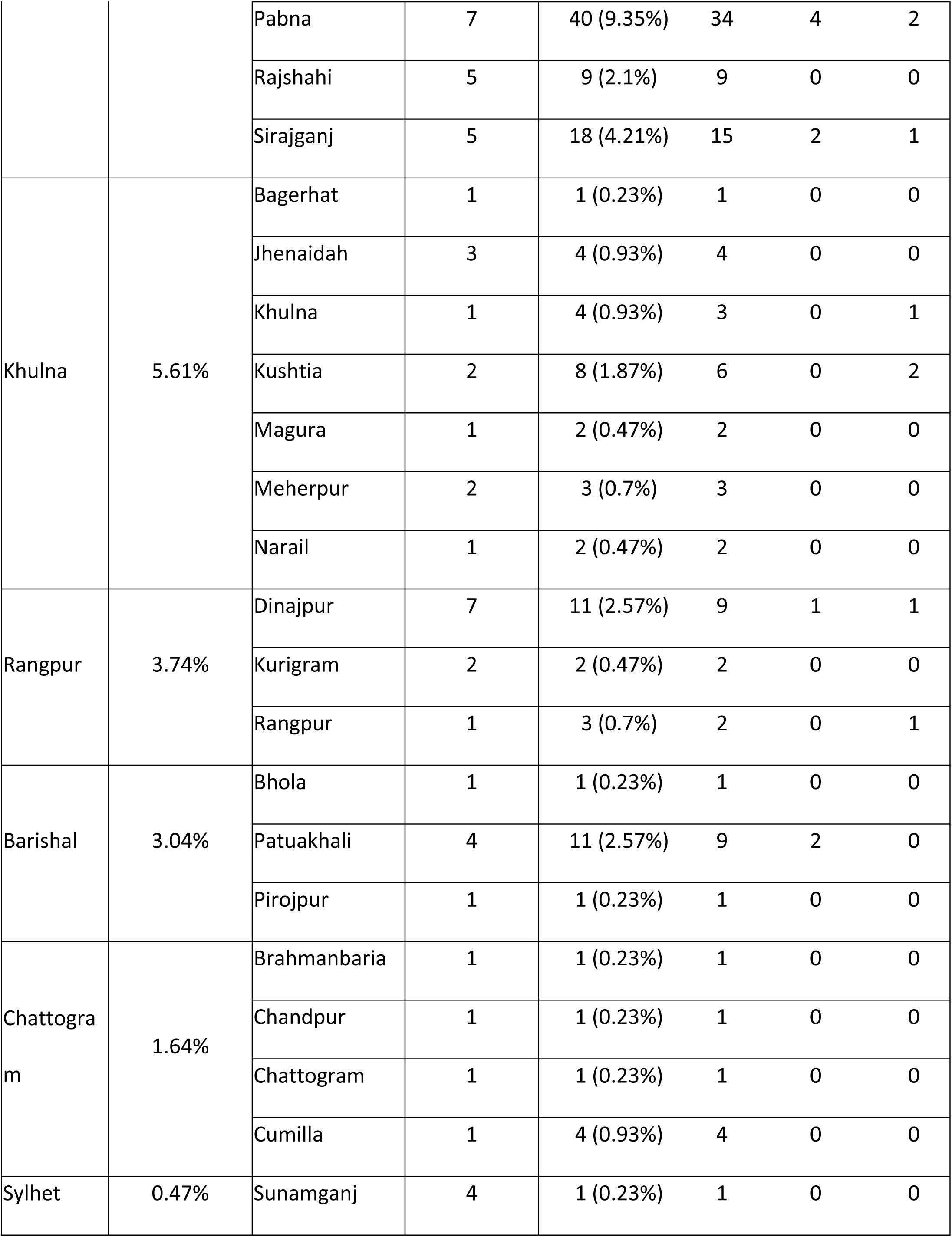
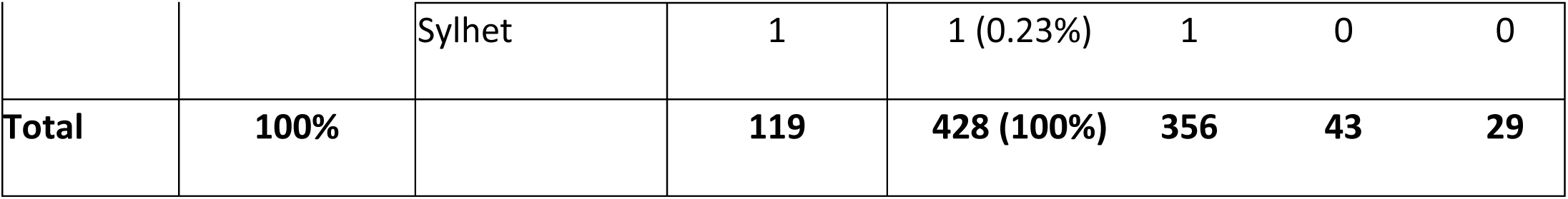
District-wise distribution of endemic Upazilas and Mouzas for VL.

**S2 Table:** Category of Mouzas with reported leishmanial infection and available sandfly data between 2017-2022.

| **Sandfly collection area (N=51)** | **No. of NKA case** | **No. of total mouza** |
| --- | --- | --- |
| Non-endemic | 0 | 9 |
| Low endemic (50 – 75 percentile) | 1 | 25 |
| Moderate endemic (90 – 95 percentile) | 2 | 7 |
| High endemic (97 percentile) | ≥3 | 10 |

NKA: new Kala Azar cases

**S3 Table:** Sandflies density with Endemic and Non-endemic HHs between 2017-2022.

| Indicators | Non-endemic<br>HH=39 | Endemic<br>HH=538 | P-value | Total<br>HH=577 |
| --- | --- | --- | --- | --- |
| **Total Sandfly, Mean ± SD** | 54.10 ± 58.27 | 8.99 ± 23.98 | <0.0001 | 12.04 ± 29.81 |
| **Total *P.argentipes*, Mean ± SD** | 40.72 ± 40.45 | 4.78 ± 18.86 | <0.0001 | 7.20 ± 22.83 |
| **Female *P.argentipes*, Mean ± SD** | 22.31 ± 23.67 | 2.49 ± 10.24 | <0.0001 | 3.83 ± 12.63 |

HH: households

